# Differential T cell clonal dynamics underlie outcomes to frontline chemoimmunotherapy in advanced gastric cancer

**DOI:** 10.1101/2025.08.26.25334455

**Authors:** Samuel J. Wright, Sarah Kang, Minae An, You Jeong Heo, Milan Parikh, Lynn Bi, Hyuk Lee, Graydon Moorehead, Nicholas Haradhvala, Sung Hee Lim, Seung Tae Kim, Gad Getz, Nir Hacohen, Jeeyun Lee, Arnav Mehta, Samuel J. Klempner, Ryan J. Park

## Abstract

The addition of aPD1 to 5-FU/platinum in advanced gastric cancer (GC) yields variable responses. To understand cooperativity between chemotherapy and immunotherapy, we previously reported a phase II trial sequentially adding pembrolizumab to 5-FU/platinum. In this study, we use single-cell RNA- and TCR-sequencing to analyze 66,813 T cells from primary tumor biopsies pre-treatment, post-chemotherapy, and post-immunotherapy in 33 patients. We observed greater abundance, persistence, and recruitment of T cells with predicted tumor-reactivity in patients with prolonged progression-free survival (slow progressors). Increased B cell abundance and predicted B cell to T cell interactions supported T cell memory and co-stimulation, providing a mechanism for increased abundance and persistence of progenitor-exhausted and tumor-reactive T cells in slow progressors. T cell clones emerging in the tumor after immunotherapy were in the blood before treatment only in slow progressors. Our study thus highlights pre-treatment and early chemotherapy-induced T cell dynamics and B cell to T cell interactions that may drive durable response to chemoimmunotherapy in GC.

## Introduction

The addition of anti-PD1/PDL1 immune checkpoint inhibitors (ICIs) to 5-fluorouracil and platinum based chemotherapy has improved survival in advanced gastric cancer (GC)^1–3^. However, the benefits are modest with a median 2-3 month gain in survival time, largely restricted to PD-L1 positive patients. Currently, we lack insights into baseline features and on-treatment adaptations that define the subgroup who derive the greatest benefit from the addition of ICIs. Analyses from the phase III Checkmate-649 trial showed no correlation between inflammatory gene signatures in bulk RNA pre-treatment biopsies and overall survival, underscoring the need for more granular approaches to dissect translational biomarkers and mechanisms^4^. We have previously identified T-cell receptor (TCR) repertoire diversity as a predictor of durable benefit in gastric cancers with microsatellite instability treated with pembrolizumab^5^. Similarly, others have shown differentiation of Tc17 (IL17+ CD8+ T-cells) toward exhausted phenotypes are permissive of tumor progression in GC^6^. However, these datasets have focused on pre-treatment samples and we lack insight into the dynamics of T cells during standard therapies.

As T-cells are the primary effectors of the anti-tumor response and target of anti-PD1 agents, we investigated T-cell clonal dynamics through sequential chemotherapy and chemoimmunotherapy and their relationship to clinical outcomes in a recently completed investigator sponsored GC trial^7^. We performed single-cell RNA-seq and single-cell TCR sequencing from serial tumor tissue biopsies and bulk TCR sequencing from serial peripheral blood samples. To account for the heterogeneity of radiographic response patterns with immunotherapy we assigned patients to clinical response categories based on progression-free survival (slow vs fast progressors^8,9^. We hypothesized that slow vs fast progressors would exhibit differences in intratumoral pre-treatment T cell composition and early chemotherapy-induced T cell evolution. Slow progressors exhibited greater abundance and persistence of GZMK+ progenitor-exhausted T cells and T cells with predicted tumor-reactivity, potentially mediated by CXCR5:CXCL13 driven recruitment of B cells that then support T cell persistence through co-stimulation and memory cytokine signaling. These findings suggest that crosstalk between germinal center-like CXCR5+ B cells and progenitor-exhausted CXCL13+ T cells sustain tumor-reactive T cell activity that underlies differential responsiveness to chemoimmunotherapy in advanced GC.

## Results

### Differential T-cell phenotypic distributions exist prior to therapy and correlate with patient outcomes

We obtained 94 primary tumor biopsies from 33 patients with treatment-naïve advanced GC. Samples were obtained pre-treatment (B), after 1 cycle of 5-FU/Platinum with trastuzumab for HER2+ tumors (F1), and after 7 cycles of pembrolizumab added to chemotherapy (F2) (**Fig. 1A**). Samples were subject to paired single-cell RNA-sequencing (scRNA-seq) and single-cell T Cell Receptor-sequencing (scTCR-seq). Additionally, patient-matched peripheral blood mononuclear cell (PBMC) samples were obtained and subjected to bulk TCR-sequencing (**Fig. 1A**).

**Figure 1.**
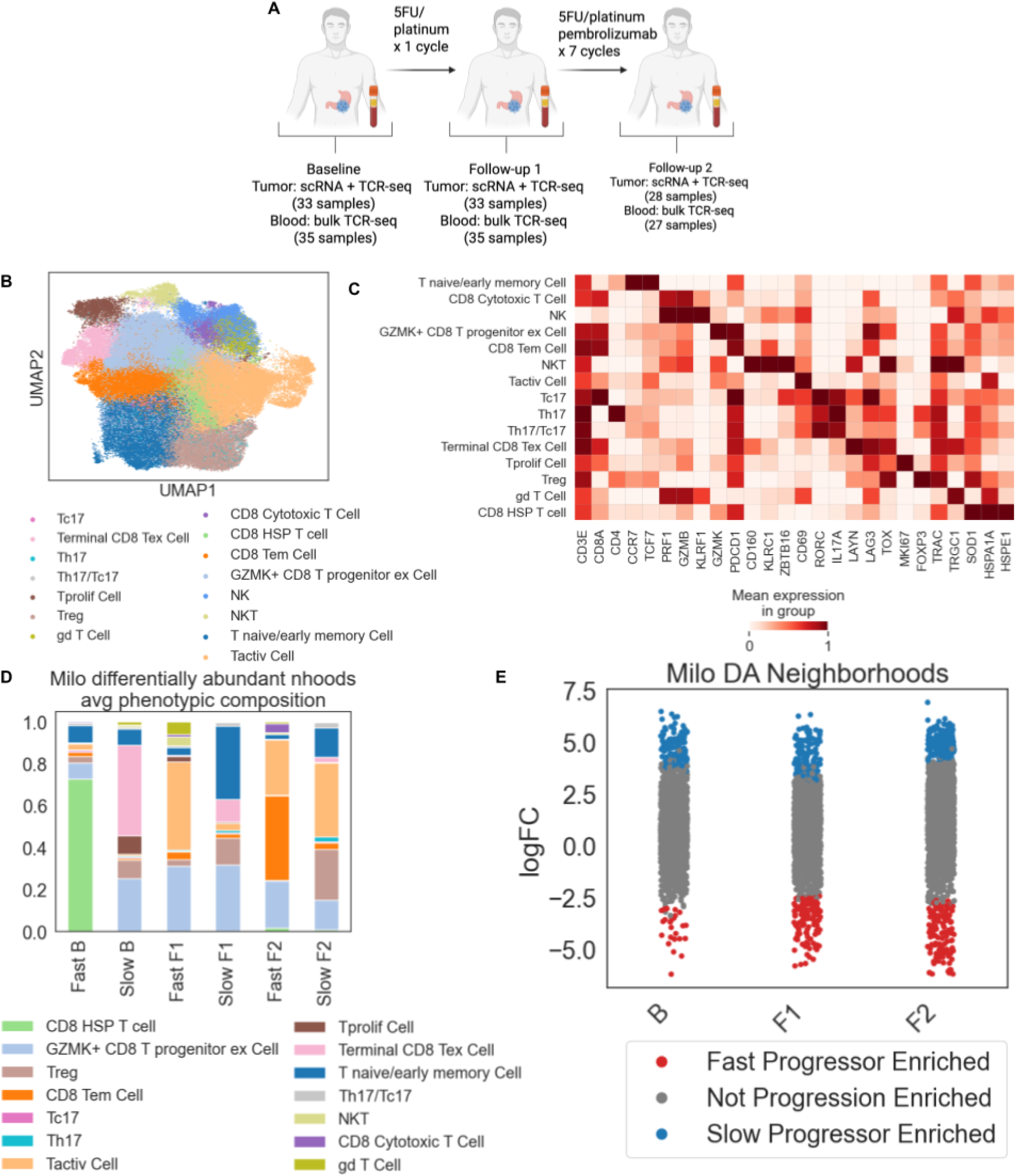
Frontline chemoimmunotherapy trial in advanced gastric cancer stratifies slow and fast progressors and identifies longitudinal transcriptomic niches (A) Diagram of sample collection from baseline, post-chemotherapy and post-chemoimmunotherapy time points, with data modalities listed next to the corresponding time points. (B) UMAP embedding of T and NK cells, annotated by subtype. (C) Average expression of canonical T cell differentiation trajectory marker genes in different T cells subsets annotated. (D) Stacked bar plot showing phenotypic composition of Milo cellular neighborhoods, averaged across neighborhoods, enriched in cells from slow or fast progressors, at each time point. e, Log fold change of milo cellular neighborhoods at each time point, colored red if enriched in cells from fast progressors and colored blue if enriched in cells from slow progressors (Spatial FDR < 0.1).

Four patients had HER2+ tumors, one patient had a EBV+ tumor, one patient had a MSI-high tumor and 29 patients had PD-L1 positive tumors. 21 patients (63.6%) achieved a best overall response of partial response (PR), 11 patients (33.3%) achieved a best overall response of stable disease (SD) and one patient (3.0%) achieved a best overall response of progressive disease (PD). Because RECIST may underestimate clinical response in single-arm immunotherapy trials we stratified patients into slow (progression-free survival (PFS) > 6 months) and fast (PFS < 6 months) progressors, with 6 months selected from median PFS in phase III trials in this patient population^1–3^. As previously described, median overall survival (OS) was not reached in slow progressors vs 263 days in fast progressors (p<0.0001).

We obtained 281,104 single cell transcriptional profiles from tumor tissues and annotated major cell types (immune, stromal, and epithelial) using a low-dimensional (UMAP) embedding, and used Leiden clustering to cluster transcriptionally similar cells in this embedding (**Fig. S1A**). We separated out clusters with PTPRC RNA expression as immune cells, and further subclustered these cells into B, Plasma, myeloid and T/NK cell phenotypes (**Fig. S1B**). We separated out a cluster expressing canonical marker genes for either or both of T and NK cells (CD3D/E/G, CD8A/B, CD4, GNLY, NKG7, etc.). Finally, we subclustered 103,894 T/NK cells from tumor and normal samples and used canonical marker gene expression across Leiden subclusters to assign T cell phenotypes (**Fig. 1B,C**). We identified 14 T cell subtypes and one NK cell subtype based on marker gene expression.

To identify differences in T cell composition between slow and fast progressors at each time point we leveraged Milo, a statistical method to identify cellular neighborhoods and compute differential abundances of neighborhoods across conditions (**Fig. 1D,E**). Pre-treatment, T cell states associated with tumor-reactivity – including terminally exhausted CD8, progenitor exhausted, and proliferating T cells – were enriched in slow progressors while HSP CD8 T cells were enriched in fast progressors pre-treatment. After 1 cycle of chemotherapy, naive/early memory T cells were newly enriched in slow progressors, suggesting chemotherapy induced new T cell recruitment to the tumor. After addition of pembrolizumab, Tregs and naive/early memory T cells were enriched in slow progressors while CD8 effector memory T cells were enriched in fast progressors, potentially reflecting short-lived, non-specific, or bystander responses. These findings suggest that differences in pre-treatment T cell composition and early chemotherapy-induced T cell remodeling shape subsequent response to PD-1 blockade.

### Clonal dynamics in slow progressors are skewed towards improved maintenance of progenitor exhausted function

We next investigated T cell differentiation trajectories and clonal dynamics across time in slow and fast progressors. Using TCR-seq, we obtained 66,813 T cell transcriptional profiles with paired alpha + beta TCR sequences (**Fig. S1C**). 2.5% of cells with TCR information were annotated as non-T cells, likely due to imprecise clustering of gene expression data, and there was not a consistent pattern in the non-T cell types across samples that had TCR information (**Fig. S2D,E**). Across samples, around 50% of cells from the scRNA-seq data were used for scTCR-seq analysis (**Fig. S2F**).

We examined clonal expansion in each T cell state across treatment time points in slow and fast progressors. Terminally exhausted CD8 T cells (Ttex) exhibited marked clonal expansion after one cycle of chemotherapy in slow but not fast progressors (**Fig. 2A**), consistent with our finding that Ttex cells are enriched in slow progressors at this timepoint (**Fig 1E**). Notably, there was no further clonal expansion in Ttex after pembrolizumab. Proliferating T cells also showed an increase in clonal expansion in slow progressors after both chemotherapy and immunotherapy, while those in fast progressors became less clonally expanded over treatment (**Fig. 2A**).

**Figure 2.**
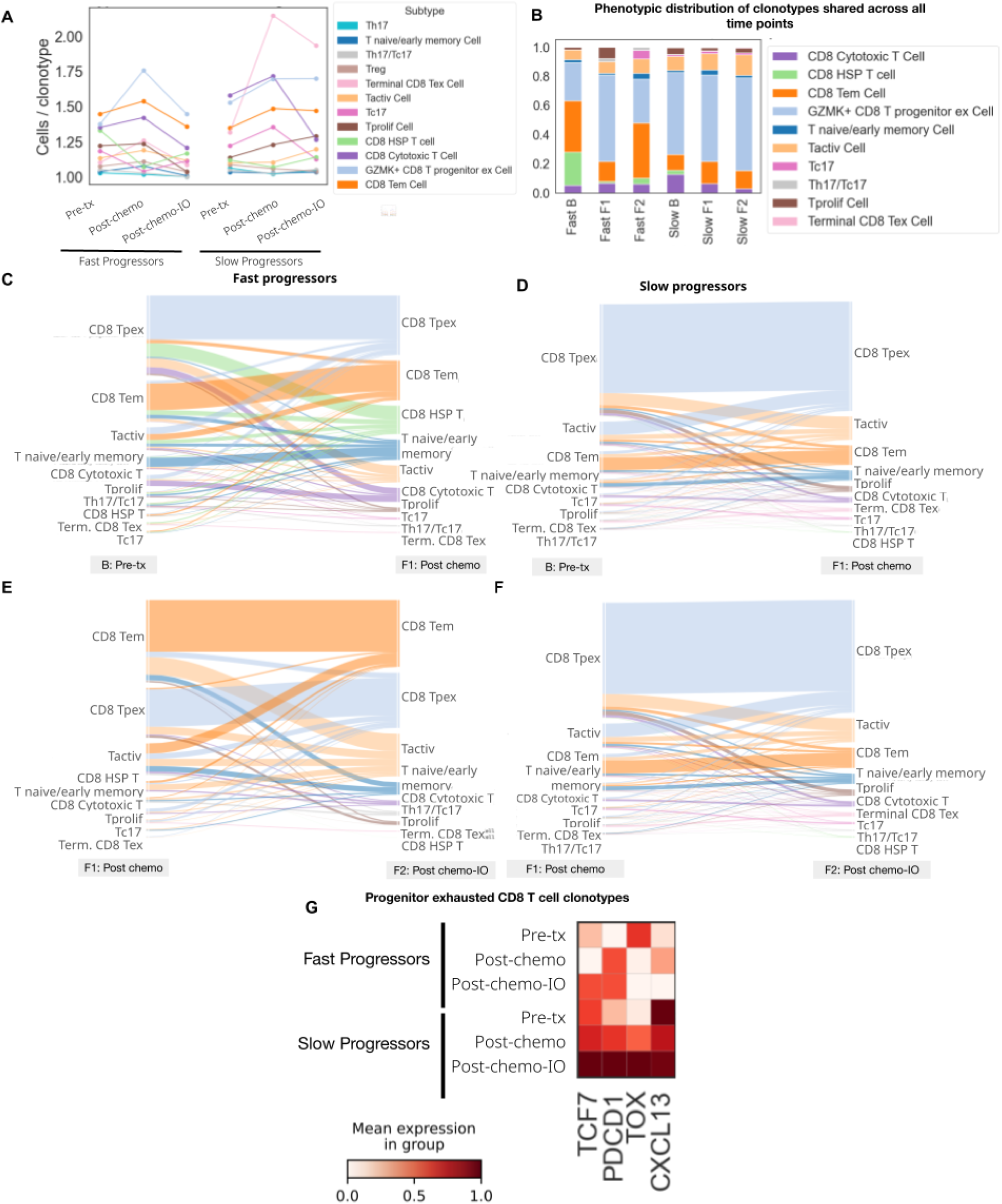
Clonal dynamics in slow progressors are skewed towards improved maintenance of progenitor exhausted function over treatment (A) Number of cells divided by the number of clonotypes within each T cell phenotype at each treatment time point in fast progressors (left) and slow progressors (right). (B) Phenotypic composition of CD8 T cell clonotypes that are shared across all time points. Each sample was randomly downsampled to 100 cells (if the sample had that many CD8 T cells) before visualizing overall phenotypic compositions. (C)-(F) Alluvial plots showing phenotypic composition of individual clonotypes shared between baseline and post-chemotherapy (C)-(D) and post-chemotherapy and post-chemo/IO (E)-(F) in fast (C) and (E) and slow (D) and (F) progressors, shown by predominant phenotype at the later (right) time point in each plot, and full proportional composition shown on earlier (left) time point in each plot. For clonotypes for which more than one phenotype was most frequent, for the purposes of visualization a predominant phenotype for that clonotype between the most frequent ones was chosen at random. (G) Average gene expression of T cell markers for stemness, exhaustion and activation at each time point in cells originating from clonotypes shared between post-chemotherapy and post-chemo/IO in fast progressors (top) and slow progressors (bottom).

We next compared the phenotypic profiles of clones found in all three time points or between pairs of treatment time points (pre-treatment to post-chemotherapy and post-chemotherapy to post-chemo/IO) (**Fig. 2B, Fig. S2A-C**). The proportions of shared clones were similar in slow and fast progressors (**Fig. S2D-E**), but shared clones were predominantly progenitor-exhausted CD8 T cells (Tpex) at all timepoints in slow progressors, while T cell states were more heterogeneous in fast progressors.

To examine T cell state changes induced by treatment, we performed TCR clonotype-based lineage tracing. Tpex and effector-memory T cells, which comprised the majority of shared clones, largely retained their state throughout treatment (**Fig. 2C-F**. In line with these findings, Tpex in slow progressors had an expression signature consistent with greater stemness, activation, and tumor-reactivity compared to fast progressors (**Fig. 2G**). In contrast, very few terminally exhausted CD8 T cells (Ttex) were present among shared clones, indicative of the limited persistence of this T cell state. Importantly, this suggests that the clonal expansion and enrichment of CD8 Ttex after chemotherapy in slow progressors (**Fig. 1E**, **Fig. 2A**) results from infiltration of new T cell clones from outside the tumor, rather than differentiation from pre-existing T cells within the tumor.

Unlike those of shared clones, the phenotypic distributions of non-shared clones were similar between slow and fast progressors (**Fig. S2F**). Similarly, the phenotypic profile (**Fig. S2G-J**) and proportion (**Fig. S2K-L**) of new clones at F1 (**Fig. S2G-H**) and F2 (**Fig. S2I-J**), while varying across patients, also did not vary significantly between progression groups. Altogether, we find that high abundance and persistence of progenitor-exhausted T cell clones throughout treatment most clearly distinguishes slow vs fast progressors and may underlie divergent response to chemoimmunotherapy.

### Putative tumor reactive clones are maintained preferentially in slow progressor

We next sought to infer tumor reactive clones as these often comprise a subset of progenitor exhausted clones^10,11^. Using a composite score for tumor reactivity based on a previously published transcriptional signature, we calculated tumor reactivity scores across CD8 cells for which we had paired RNA and TCR data (**Fig. 3A**) ^12^. We annotated putative tumor reactive clones by clonotypes with at least one cell surpassing a pre-determined threshold (**Fig. S3A-J**).

**Figure 3.**
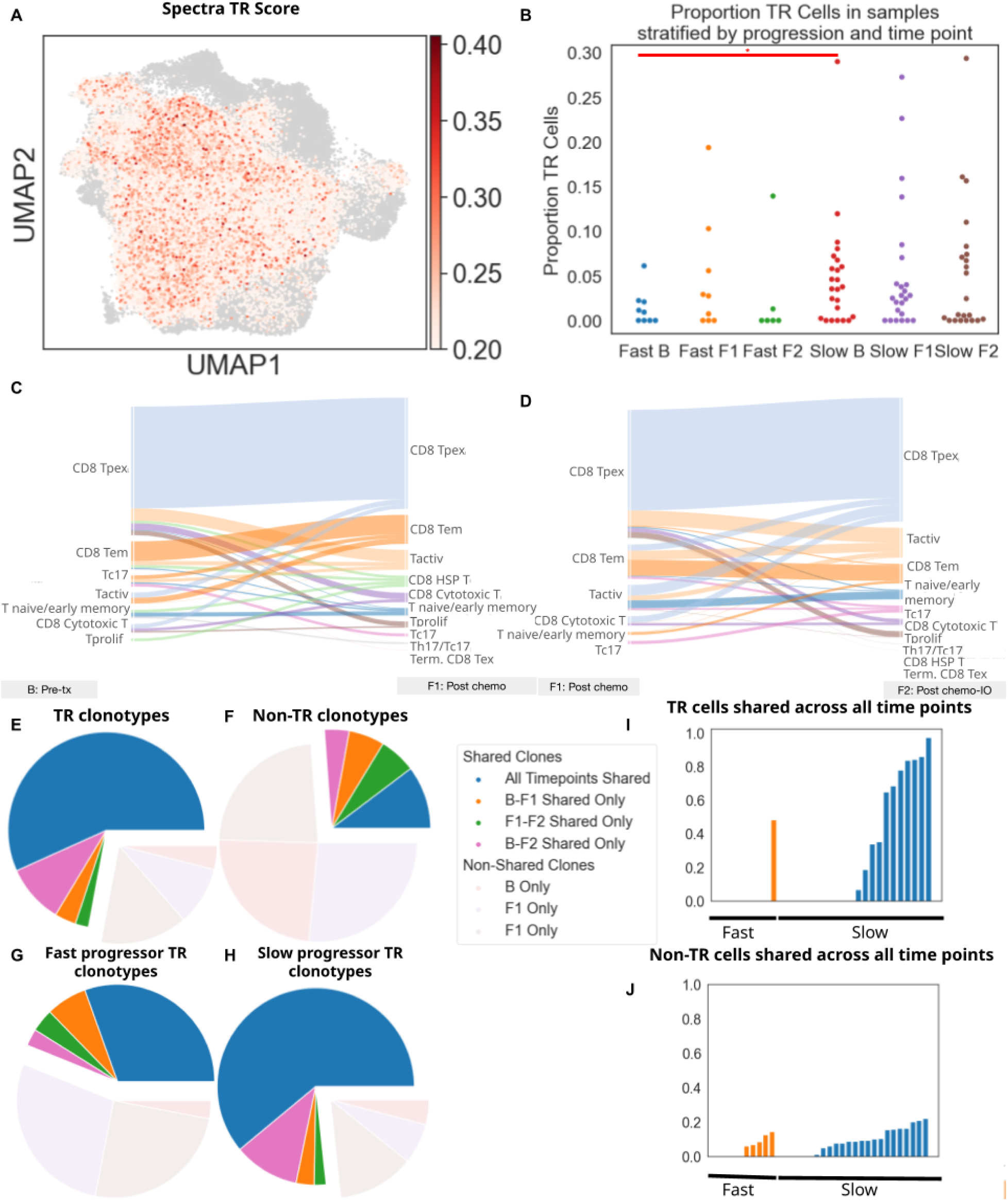
Putative tumor reactive clones are maintained preferentially in slow progressor tumors compared to fast progressors. (A) UMAP embedding, colored by tumor reactivity scores of CD8 T cells with TCRs. (B) Swarm plot showing proportion of cells among CD8 T cells with TCRs that are putatively tumor reactive in each sample, stratified by progression status and time point. Wilcoxon rank sum test, * : p < 0.05. (C)-(D) Alluvial plots showing phenotypic composition of individual clonotypes shared between baseline and post-chemotherapy (C) and post-chemotherapy and post-chemo/IO (D) among putative tumor reactive clonotypes. (E)-(H) Pie charts showing time points across which putative tumor reactive (E) and non-tumor reactive (F), and putative tumor reactive clonotypes in slow (G) and fast (H) progressors are present. Clonotypes shared between at least two time points are shaded darker than non-shared clonotypes. (I)-(J) Proportion of putative tumor reactive out of all tumor reactive cells (I) and non-tumor reactive, out of all non-tumor reactive cells (J) clones in each patient in fast (left, orange) and slow (right, blue) progressors.

We first noted that slow progressors have a higher proportion of putative tumor reactive cells per sample among CD8 T cells pre-treatment, and that these proportions seem to be better maintained throughout treatment than in fast progressors (**Fig. 3B**). Tumor reactive clones are phenotypically skewed towards a progenitor exhausted CD8 T cell phenotype and do not undergo substantial state transitions (**Fig. 3C,D, Fig. S3K-O**). Tumor reactive clones persisted in the tumor through two or more treatment time points while non-tumor reactive clones largely did not (**Fig. 3E,F**). Additionally, tumor-reactive clones predominantly persisted across multiple time points in slow progressors but were significantly less persistent in fast progressors and non-tumor-reactive clones were not persistent in either group (**Fig. 3G,J).** To examine the persistence of non-tumor-reactive T cells through another approach, we matched viral-reactive and bacterial-reactive CDR3 sequences from public databases to our data. Clonotypes matching these sequences were present in gastric tumors to varying degrees but were minimally persistent across timepoints in both slow and fast progressors (**Fig S4**).

### Circulating clonal frequencies precede infiltration of progenitor exhausted CD8 T cells and correlate with tumor microenvironment features

To investigate blood to tumor trafficking, we performed bulk TCR-seq of peripheral blood mononuclear cells (PBMCs) time-matched to tumor biopsies (**Fig. S5A-B**). In slow but not fast progressors, TCR clonotypes emerging in the tumor after immunotherapy pre-existed in the blood prior to treatment and after chemotherapy (**Fig. 4A**). For these TCRs which emerged after immunotherapy and pre-existed in the blood prior to treatment and after chemotherapy, we investigated the state of T cells belonging to these clonotypes. In slow but not fast progressors, these T cells were enriched in a progenitor exhausted state (**Fig. 4B, S5C**). Additionally, the abundance of progenitor exhausted cells with these TCRs was directly correlated with the frequency of the same TCRs in the blood pre-treatment and post-chemotherapy (**Fig. 4C).** Clonotypes shared between blood and tumor at other timepoint pairs were also predominantly Tpex in the tumor (**Fig. S5D-E**).

**Figure 4.**
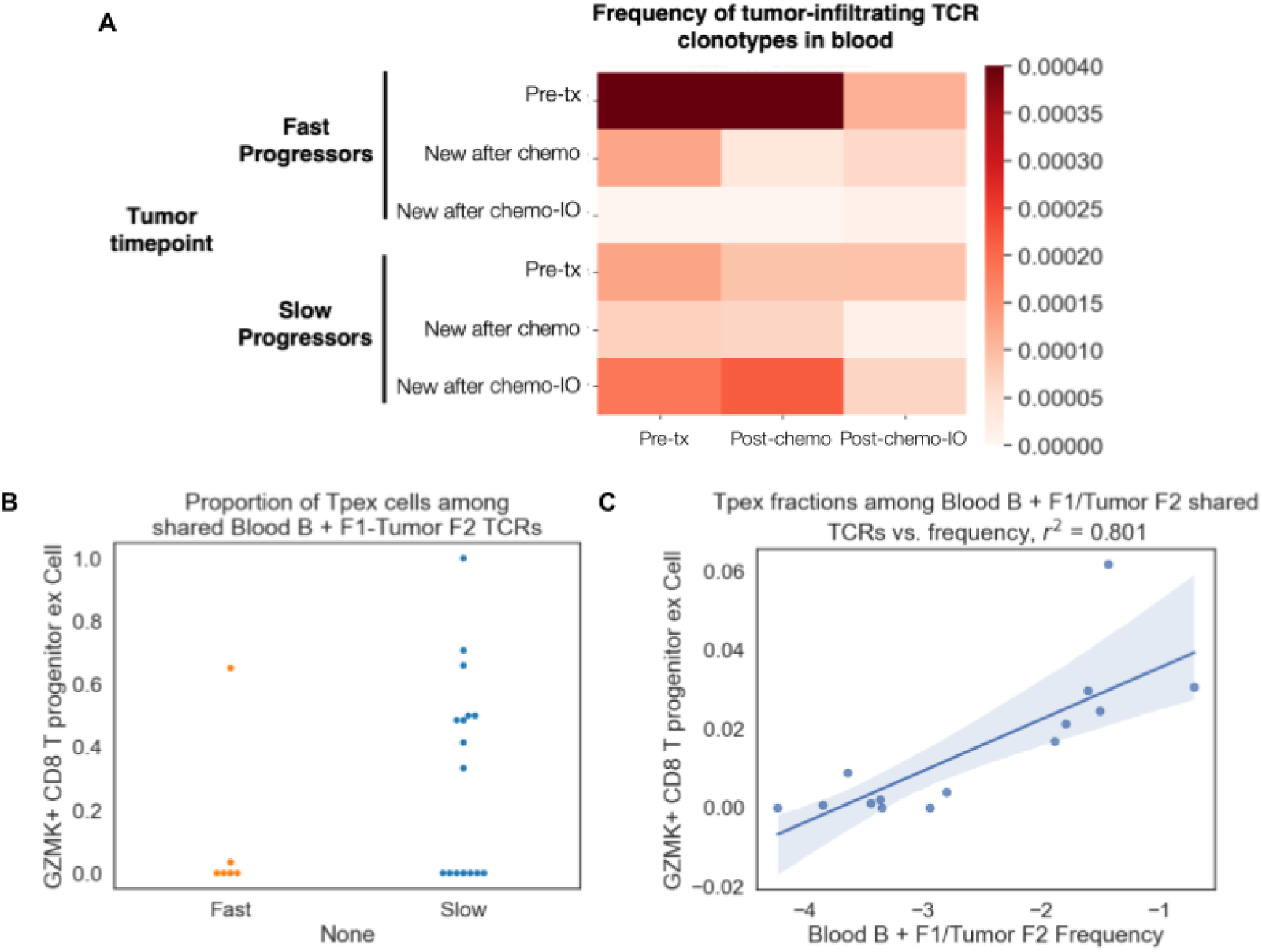
Circulating clonal frequencies precede infiltration of progenitor exhausted CD8 T cells and correlate with cellular orchestration in the tumor microenvironment (A) Total blood frequencies of circulating TCR clones that are shared with tumor clones present in pre-treatment tumor, those new in post-chemotherapy tumors and those new in post-chemo/IO tumors in fast (top) and slow (bottom) progressors. (B) Proportion of progenitor exhausted CD8 T cells out of all T cells with TCRs post-chemo/IO shared with TCRs in the blood at either pre-chemo or post-chemo in fast (left) and slow (right) progressors. Wilcoxon rank sum test, p = 0.302. (C) Scatterplot with linear regression of proportion of progenitor exhausted CD8 T cells with TCRs post-chemo/IO shared with TCRs in the blood at either pre-chemo or post-chemo, divided by total number of T cells, against combined tumor post-chemo/IO shared blood pre-chemo and post-chemo TCR frequencies (log scale).

### B cell dynamics correlate with response and co-correlate with shared progenitor exhausted T cell clones

Finally, we observed marked differences in B cell abundance in slow vs fast progressors (**Fig. 5A**). In slow progressors, B cell abundance increased throughout treatment, while in fast progressors B cell abundance did not rise after chemotherapy and were nearly absent in all patients. Additionally, cDC1 (**Fig. S6A**) and cDC2 (**Fig. S6B**) cells showed similar trends between slow and fast progressors over time, albeit to a lesser extent than B cells, suggesting that these cells may be contributing to the increased tumor-reactivity and activation of T cells that we have observed.

**Figure 5.**
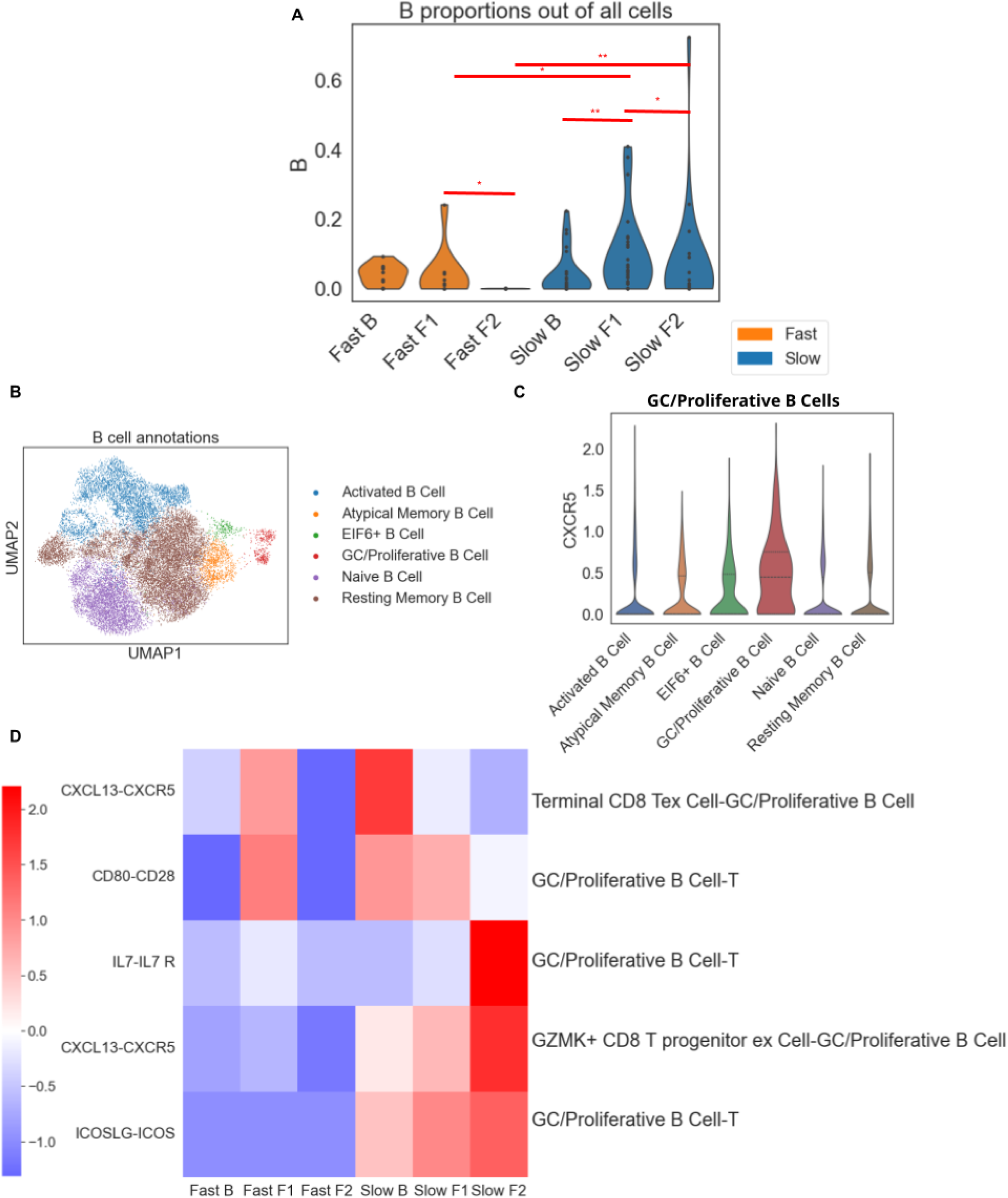
B cell dynamics correlate with response and co-correlate with shared progenitor exhausted T cell. (A) Proportions of B cells in each patient out of all cells in each sample across sampling time points. Wilcoxon rank sum test, * : p < 0.05, ** : p < 0.01. (B) UMAP clustering and annotation of B cells across patients. (C) Distribution of CXCR5 expression across B cell subsets. (D) Heatmap showing relative interaction strengths between B and T cell subsets across different interactions, stratified by time point and progression.

We identified 6 B cell subsets using canonical marker genes (**Fig. 5B**)^13^. GC/Proliferative B cells showed the highest expression of CXCR5 (**Fig. 5C**), the receptor for CXCL13^12^ which is strongly associated with tumor-reactive T cells. Similar to overall B cells, GC/Proliferative B cell proportions rose throughout treatment in slow progressors, consistent with ongoing recruitment by CXCL13+ T cells, while in fast progressors they rose slightly post-chemo before dropping post-chemo/IO (**Fig. S6C**).

To identify TME features associated with B cell phenotypes at each treatment timepoint, we correlated B cell subtype proportions with those of the cell types across samples. Pre-treatment (**Fig. S6D**), GC/Proliferative B cells were correlated with proliferating and terminally exhausted CD8 T cells, which may relate to the potential clonal expansion trends we observed in **Fig. 2A**. Post-chemotherapy (**Fig. S6E**), GC/Proliferative B cells again correlated with terminally exhausted CD8 T cells and also with cDC1. Post-chemo/IO (**Fig. S6F**), GC/Proliferative B cells correlated with cytotoxic CD8 T cells, potentially reflecting B:T interactions that promote anti-tumor T cell activity To investigate potential B:T interactions, we employed CellPhoneDB^14^ (**Fig. S6G**). We examined B:T interaction strength for selected receptor:ligand pairs stratified by progression group and across time (**Fig. 5D**). CXCL13-CXCR5 interaction strength was higher in slow progressors, likely mediating ongoing B cell recruitment to the tumor and specifically to Tpex cells. ICOS-ICOSLG and CD80-CD28 interaction strengths were higher in slow progressors, potentially mediating improved costimulation and therefore activation of T cells. IL7-IL7 receptor strength was higher in slow progressors post-chemo/IO and is an important mediator of T cell memory and persistence.. Together, these results suggest a central role for B:T communication in maintaining and amplifying T cell activity, as CXCL13 from T cells recruits CXCR5+ B cells, and B cells in turn provide co-stimulation and IL7 signaling to T cells to sustain their effector functions.

## Discussion

The development and maintenance of an anti-tumor immune response represents a complex series of multicellular interactions, and has therapeutically focused on T-cell functionality^15^. While there is broad clinical adoption of anti-PD-1 with 5FU/platinum agents in gastroesophageal cancers, the absolute benefit remains modest and the remodeling of the intratumoral T cell repertoire during therapy remains poorly understood due to a lack of longitudinal tissue sampling. To address this and identify dynamics associated with durable clinical responses, we employed serial primary tumor biopsies and paired peripheral blood analyses from a sequential chemoimmunotherapy trial to separately analyze pre-treatment tumor characteristics, early chemotherapy-induced remodeling, and chemoimmunotherapy-induced remodeling. By integrating single-cell RNA- and TCR-sequencing, we identify a coordinated program of T cell persistence and B cell support that distinguishes gastric cancer patients with prolonged clinical benefit to chemoimmunotherapy (**Figure 6**).

**Figure 6.**
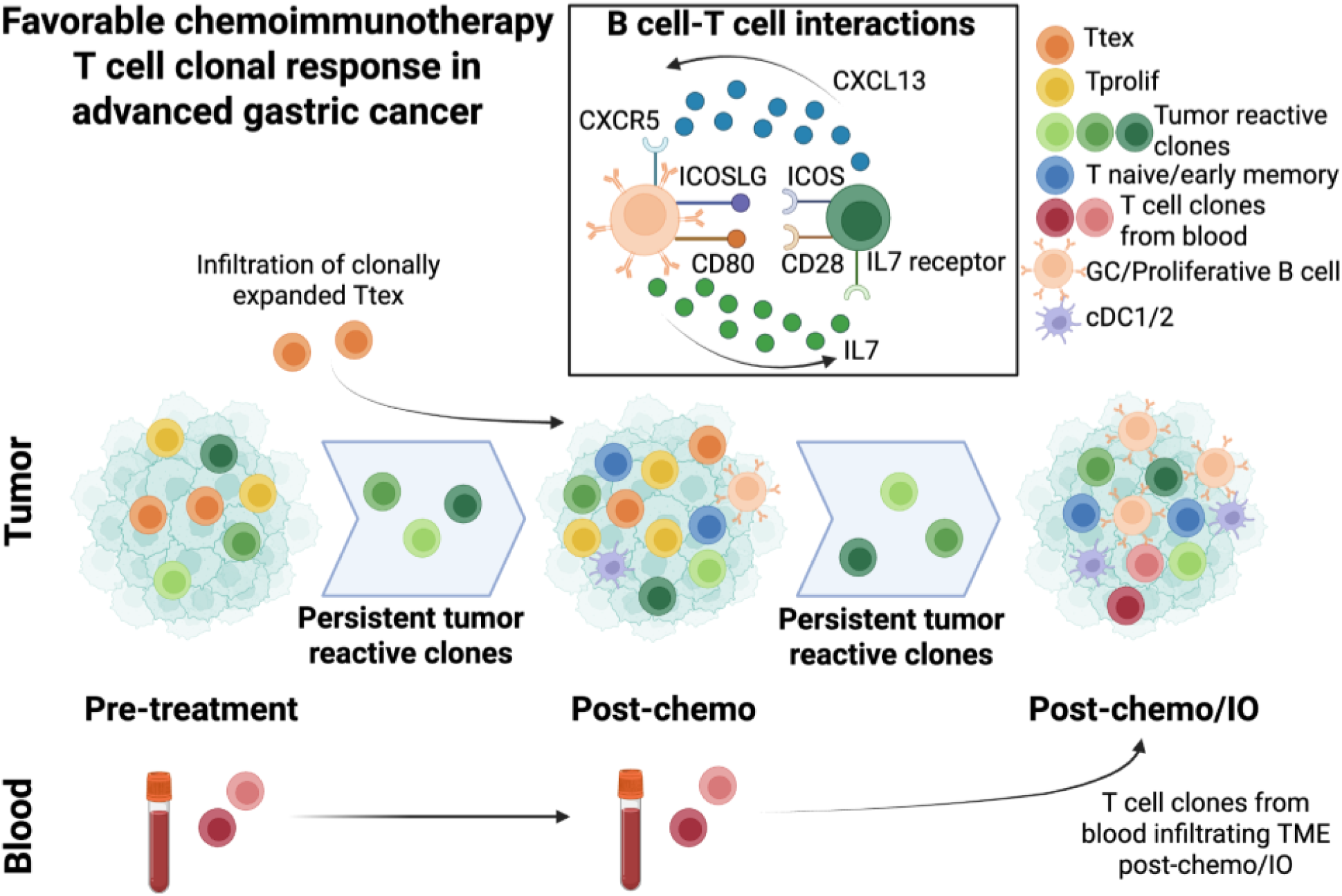
Summary diagram detailing findings from this study related to T cell clonal response in slow progressors, including persistence of tumor reactive/progenitor exhausted T cell clones in the tumor throughout treatment, infiltration of terminally exhausted clones post-chemotherapy, detection of T cell clones in the blood pre-treatment and post-chemotherapy that appear in the tumor post-chemo/IO, and interactions between T cells and B cells.

A key finding is the preferential enrichment and persistence of progenitor-exhausted (Tpex) CD8+ T cells in slow progressors. Tpex cells in slow progressors exhibited greater stemness and tumor-reactivity, persisted in the tumor throughout treatment, and preferentially shared common clonotypes with circulating T cells. In contrast, fast progressors exhibited a more heterogeneous T cell repertoire with lower abundance and persistence of tumor-reactive clones. Importantly, these distinguishing features were already present after one cycle of chemotherapy, suggesting that this early remodeling may set the stage for effective aPD-1 therapy.

Another important observation is that, in slow but not fast progressors, intratumoral Tpex cells arise from T cell clones that are already circulating in the blood prior to treatment. The frequency of pre-existing circulating T cell clones that infiltrate into the tumor after chemoIO was correlated with a more immunologically active tumor microenvironment including higher dendritic cell abundance and more proliferating T cells after chemotherapy. Our results also highlight a potentially central role for CXCR5+ germinal center-like B cells in supporting tumor-reactive T cell persistence and activation. Our model is that CXCL13+ tumor-reactive T cells recruit CXCR5+ B cells to the tumor, and these B cells in turn sustain and amplify these T cells through ICOSLG and CD80 costimulation and IL7 signaling.

Taken together, these findings help define a coordinated immune ecosystem in responsive tumors characterized by (1) pre-existing tumor-reactive progenitor exhausted T cells, (2) early chemotherapy-induced recruitment and proliferation of these cells, and (3) B cell–mediated support that promotes T cell persistence and anti-tumor activity. These insights extend previous studies which have largely focused on pre-treatment biopsies, revealing dynamic immune remodeling events that are key to therapeutic efficacy.

Our findings align with and extend a growing body of literature underscoring the central role of progenitor exhausted (Tpex) CD8+ T cells in mediating durable responses to immune checkpoint blockade. These act as a reservoir for fresh T cells and are responsible for the proliferative burst following PD-1 therapy^16,17^. While prior studies have largely focused on Tpex abundance at a single timepoint, our dense serial sampling reveals the importance of intratumoral persistence and phenotypic stability of Tpex and tumor-reactive TCR clonotypes in patients with good clinical response. These results add important nuance to existing literature that has emphasized the importance of newly infiltrating T cell clones over pre-existing intratumoral clones in aPD1 response and highlight the dual importance of persistent Tpex clones marked by phenotypic stability, as well as early chemotherapy-induced recruitment of new T cell clones that exhibit a clonally expanded, terminally exhausted phenotype^18,19^.

In parallel, enrichment of B-cells has been described in favorable response to aPD1 and the roles for B-cells in supporting T-cell responses are well described^20–23^. We find that B cell abundance closely tracks with Tpex persistence and highlight reciprocal B and T interactions as a potential mechanism for these findings.

It is important to recognize that the analysis conducted here should be considered exploratory and the primary endpoint of the clinical trial was objective response rate, with correlative associations serving as key secondary objectives. Functional testing for predicted tumor-reactive TCR sequences and spatial analyses of B andT interactions will be necessary to confirm our hypotheses. Finally, validation of our findings in a larger patient cohort will be essential to confirm generalizability of our findings.

Our findings point to several avenues for subsequent study, focused on stimulating and maintaining the phenotypic distribution of putative anti-tumor TCRs. Small studies in GI cancers have highlighted the potential for the use of putative neoantigen-selected T-cells, although overall clinical activity remains limited^24,25^. In future work we plan to leverage the WES and RNA seq data from this trial to nominate neoantigens for functional testing, however, we recognize that in current pipelines only between 6-10% of predicted neoantigens are actually recognized by T-cells ^7,25,26^. Importantly, our work suggests that tumor microenvironmental features, particularly B and T cell interactions, may play a role in early remodeling the TME to potentiate durable clinical benefit with immunotherapy. Further efforts to identify baseline and treatment-induced spatial features that underscore differential T cell abundance in patients that receive benefit are needed. As we uncover mechanisms underlying anti-tumor TCR persistence, as a field, we need to continue to develop gastric cancer model systems that are relevant for functional testing of immune pathways observed in patients.

In summary, we provide an unprecedented time-resolved dataset and analysis of T-cell dynamics during frontline chemoimmunotherapy in gastric cancers. Our data points to differential T-cell state flux between patients benefitting from 5FU/platinum and pembrolizumab with the notable ability to support persistence and expansion of putative tumor reactive progenitor exhausted CD8 populations in responding patients.

## Acknowledgments

We would like to thank funding support from the NIH/NCI Gastrointestinal Cancer SPORE P50 (AM, SJK), Doris Duke Charitable Foundation Physician Scientist Fellowship (AM) and DF/HCC K12 (K12CA087723) Paul Calabresi Award for Clinical Oncology (AM), Prostate Cancer Foundation Young Investigator Award (RJP), the Torrey Coast Foundation (GG, SJK, RP) and the Gastric Cancer Convergence Team administered by the American Association for Cancer Research (GG, SJK, RP), and a grant of the Korea Health Technology R&D Project through the Korea Health Industry Development Institute (KHIDI), funded by the Ministry of Health & Welfare, Republic of Korea (grant number : RS-2024-00437038 to JL, SJK).

## Methods

### Patients and study design

The design and primary outcome from this single arm phase II investigator sponsored trial are previously reported by our group ^7^. Briefly, patients with locally advanced, unresectable or metastatic gastric cancer with no prior therapy were enrolled. All HER2 negative patients received capecitabine with oxaliplatin for one cycle and then pembrolizumab was added to subsequent cycles. If the patient was HER2 positive then they received capecitabine with cisplatin and trastuzumab for one cycle and then pembrolizumab for all subsequent cycles. The complete inclusion and exclusion criteria and trial details are included as supplementary material. The trial protocol was approved by the Samsung Medical Center IRB (IRB # 2019-11-089) and registered on clinicaltrials.gov (NCT04249739). All patients provided written consent prior to enrollment and the trial was conducted in accordance with the Declaration of Helsinki and the Guidelines for Good Clinical Practice.

### Patient sample processing

Tissue samples were collected from the patients who had biopsies one day prior to cycle 1, one day before cycle 2 and 1 day before cycle 7. All tissue samples were from the primary gastric tumor which was endoscopically mapped so the same region of the tumor was sampled at each biopsy. Matched peripheral blood was collected prior to initiation of treatment and at serial timepoints aligned with tumor samples as described in the protocol. For tissue samples, if tumor purity was estimated to be more than 40% after pathological reviews, tumor DNA and RNA were extracted by using a QIAamp Mini Kit (Qiagen, Hilden, Germany) according to the manufactures’ instructions for exome and transcriptome sequencing. Concentration, 260/280 and 260/230nm ratios were measured with ND1000 spectrophotometer (Nanodrop Technologies, Thermo-Fisher Scientific) and then DNA/RNA was quantified using a Qubit fluorometer (Life Technologies).

### scRNA-seq data preprocessing

We aligned scRNA-seq reads to the GRCh38 human genome reference and quantified them using a Cellranger (version 5.0). DoubletDetection was used to filter out doublets^27^. Cells with high mitochondrial read proportion (>20%) and more than 6,000 detected genes were filtered out. Data from each sample was normalized and scaled using the Scanpy (v1.9.8) pp.log1p function and principal components were calculated using the Scanpy (v1.9.8) tl.pca function. Uniform Manifold Approximation and Projection (UMAP) was calculated on the data using the by calculating nearest principal component neighbors with the Scanpy (v1.9.8) pp.neighbors function and embedding using Scanpy (v1.9.8) tl.umap function. Leiden clusters were determined using the Scanpy (v1.9.8) tl.leiden function. Differentially expressed genes determined using Mast, in addition to canonical marker genes, were used to annotate clusters^28^. Clusters were initially annotated as epithelial, immune and stromal before each of these populations was subclustered for further annotation. In some subclusterings where batch effects were evident in the clustering, Harmony was employed to generate a batch-corrected dimensionally-reduced space using Scanpy’s (v1.9.8) external.pp.harmony_integrate function.

### Differential abundance analysis

Milopy (v0.1.1) was used to calculate T cell neighborhoods that were enriched in cells from either fast or slow progressors at each time point. First, cells from a given time point were selected, dimensionally-reduced spaces were re-embedded using the same process as described above. Cell neighborhoods were calculated and calculated, and differential abundances were calculated using the milopy (v0.1.1) make_nhoods, count_nhoods and DA_nhoods functions, respectively, as described in the available tutorial online: https://milopy.readthedocs.io/en/latest/milopy_example.html. Cell neighborhoods who were composed of a proportion of a single annotated cell type below 60% were considered “Mixed” and excluded from visualization. Using milopy’s (v0.1.1) statistical framework, neighborhoods with a Spatial FDR below 0.1 were considered differentially abundant in either cells from slow or fast progressors.

### scTCR-seq data preprocessing

We aligned scTCR-seq reads to the GrCh38 human genome reference and quantified them using Cellranger (version 6.0.1) for using ‘vdj’ as the input data type. Scirpy (v0.12.0) was used to integrate scTCR reads for each sample with its corresponding scRNA sample. Clonotypes were determined based on distances between nucleotide or amino acid sequences using the Scirpy (v0.12.0) pp.ir_dist and tl.define_clonotypes function. Cell barcodes without matching scRNA-seq data, without non-null alpha or beta CDR3 sequences and/or matching a scRNA-seq barcode annotated as a cell type other than T cell were filtered out from downstream analysis.

### Cells per clonotype

For each T cell phenotype at each time point, within each sample the number of cells belonging to that phenotype was divided by the number of clonotypes present across that cell phenotype. At each time point, the cells per clonotype value was averaged across slow progressors and fast progressors separately.

### Downsampling for phenotypic distribution visualization

For stacked bar plots indicated in the results section, cells in each sample were randomly downsampled to 100 T cells with scTCR data per sample (unless the sample had less than 100 T cells with scTCR data per sample, in which case the total number of cells was used for that sample), before stratifying samples by time point and progression status and finding the proportion of subsampled cells in those samples belonging to each phenotype.

### Alluvial plot visualizations

An alluvial plot function (https://github.com/vinsburg/alluvial_diagram) was used for visualizing phenotypic transitions experienced by clonotypes between treatment time points. Plots were made such that the clonotype’s predominant phenotype is shown on the right of the plot and its per cell composition is shown on the left of the plot. For clonotypes whose most frequent phenotype was not a single cell state, one phenotype from the most frequent ones at that time point was chosen at random.

The Plotly (v5.22.0) express.parallel_categories function was used to depict the predominant state of clonotypes at each time point for clonotypes shared across all three treatment time points. Clonotypes whose most frequent phenotype was not a single cell state at a given time point were labeled as “Mixed” at that particular time point.

### Bulk TCR preprocessing

The analysis pipeline described in Li et al., 2019 was used to obtain bulk TCR CDR3 sequences found in blood samples ^29^.

### Blood-tumor TCR sharing

Shared clonotypes were determined based on beta chain CDR3 sequences that appeared in both the tumor scTCR-seq data and PBMC bulk TCR-seq data. The sum of TCR frequencies in the blood that matched with the tumor was used to determine the total shared blood TCR frequency of blood-tumor shared clones in circulation.

### Random forest regression

Random forest regression was performed using the scikit-learn (v1.3.2) ensemble.RandomForestRegression function with default hyperparameters.

### Linear regression

Linear regressions were performed using the Scipy (v1.10.1) linregress function to find relationships between continuous variables.

### Public clonotypes

TCRdb (https://guolab.wchscu.cn/TCRdb/) was used to find bacterial- and viral-reactive TCR sequences^30^. Beta chain CDR3 sequences annotated to be reactive to either bacteria or viruses were matched with those in tumor data to annotate bacterial or viral TCR clonotypes, respectively.

### Clonotype-by-clonotype phenotypic change plots

For clonotypes shared between baseline and post-chemotherapy or between post-chemotherapy and post-chemo/IO, clonotypes were stratified by progression status first and then by patient ID. Phenotypic proportions per clonotype were subtracted between the succeeding time point and preceding time point, hierarchically clustered, and plotted using the pandas (v1.5.3) plot.bar function such that phenotypes that were more frequent at the preceding time point are shown below the y=0 line and those that were more frequent at the succeeding time point are shown above the y=0 line. In other words, for clonotypes whose phenotypic composition remained the same from the preceding to succeeding time point, these clonotypes are shown as ‘blank’ or white bars on this plot.

### Inference of putative tumor reactive clonotypes

We applied a gene signature associated with CD8+ T cell tumor-reactivity derived through supervised factorization of a scRNA-seq dataset from an anti-PDL1 treated cohort with non-metastatic breast cancer^12,31^. To benchmark the tumor-reactivity signature, we utilized a scRNA-seq dataset with paired scTCR-seq of CD8+ T cells from a cohort of melanoma patients in which anti-tumor specificity had been experimentally validated and annotated^32^. A scoring threshold was established to identify tumor-reactive cells by generating an AUROC curve. Given that all cells of a T cell clonotype share antigen specificity, a clonotype was predicted to be tumor-reactive if at least one cell scored above the specified threshold. UCell was used for gene signature scoring with default parameters 4^33^. In brief, UCell calculates a normalized Mann-Whitney U statistic ranging from 0 to 1, based on the ranked expression of genes within each cell. We selected a scoring threshold of 0.285 which minimized the false positive rate (0.347) and maximized the true positive rate (0.837). This scoring strategy and threshold were subsequently applied to all T cells in our paired scRNA-seq and scTCR-seq dataset to identify putative tumor-reactive T cells.

### Covariation analysis of tumor microenvironment cell type proportions

Separately for each time point, we took the proportion of cell subtypes out of their parent subtype (B cell subtypes out of total B cells, T cell subtypes out of total T cells, stromal cell subtypes out of all stromal cells, myeloid cell subtypes out of total myeloid cells, NK cells out of total cells, epithelial cells out of total cells and plasma cell out of total cells) along with the proportion of cells in each sample that were putatively tumor reactive. We then calculated pairwise correlations between features using the pandas (v1.5.3) corr() function and hierarchically clustered their abundances with the seaborn (v0.13.2) clustermap function.

### Scoring cell-cell interactions by progression and time point groups

Interactions identified as appearing significantly between B cell and T cells were selected to compare based on their relative strengths in slow and fast progressor at each treatment time point. B and T cells were subsetted into each of these progression/time point categories. For each B and T cell subset, average expression values for the ligand and receptor gene for each interaction were taken. If the receptor involved a receptor complex, average gene expression values were taken for genes corresponding to each of the components. For each cell subtype combination between B and T cells possible, the minimum between the average ligand expression in the first cell type and average receptor component(s) expression in the second cell type was taken to determine the relative interaction strength in either direction.

### CellPhoneDB

CellPhoneDB (v5.0.0) was used to infer cell-cell interactions between cell types in the tumor, as described in the tutorial available online: https://github.com/ventolab/CellphoneDB ^14^.

### Data availability

Single-cell RNA sequencing data from the patient cohort are deposited in the European Nucleotide Archive (ENA) under accession PRJEB60680 and these data will be provided upon reasonable written request for academic use and within the limitations of the clinical trial informed consent and general data protection regulations.

## Supplemental Figure Legends

**Figure S1.**
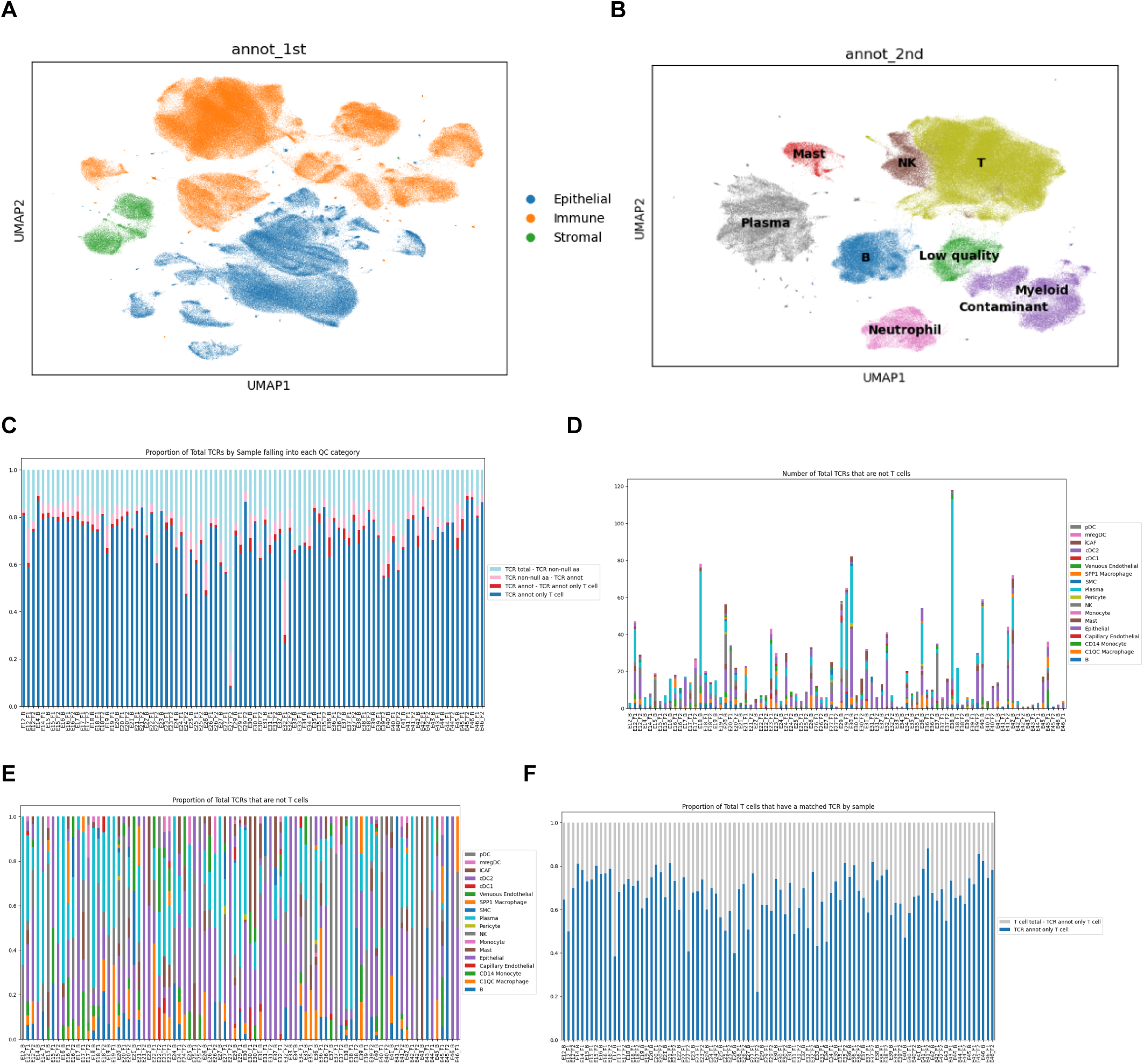
(A) UMAP embedding of all cells collected from tumor and normal samples, annotated as either epithelial, stromal or immune based on canonical marker genes. (B) UMAP embedding of immune cells from tumor and normal samples, annotated using canonical marker genes. (C) Proportion of each sample that has single-cell TCRs with a T cell annotation in the single-cell RNA-sequencing data and a non-null alpha and beta CDR3 sequence (blue), a non-T cell annotation and a non-null alpha and beta CDR3 sequence (red), a matching single-cell barcode but a null alpha and/or beta CDR3 sequence (pink) and no matching single-cell RNA-sequencing barcode (light blue). (D)-(E) Number (D) and proportion (E) of non-T cell annotation matching a cell barcode with a TCR in each sample. (F) Proportion of cells in each single-cell RNA-sequencing sample with (blue) and without (grey) matching TCR-sequencing information data.

**Figure S2.**
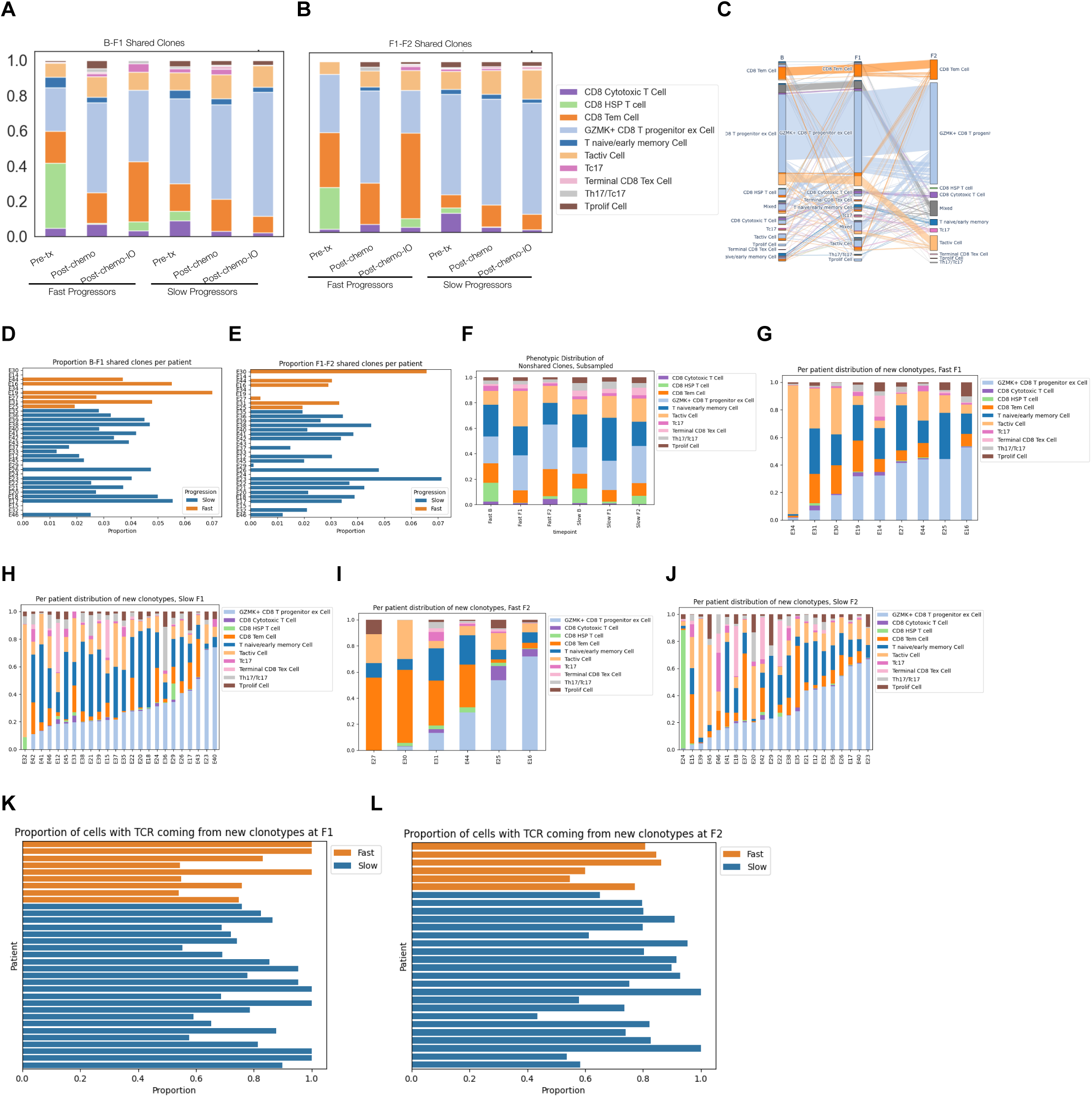
(A)-(B) Phenotypic composition of CD8 T cell clonotypes that are shared between baseline and post-chemotherapy (A) and post-chemotherapy and post-chemo/IO (B) time points at each time point. Each sample was randomly downsampled to 100 cells (if the sample had that many CD8 T cells) before visualizing overall phenotypic compositions. (C) Alluvial plot showing the predominant phenotype of clonotypes shared across all three treatment time points. Clonotypes that are not predominantly a single phenotype at a given time point were labeled as ‘Mixed’. (D)-(E) Proportion of clones shared between baseline and post-chemotherapy (D) and post-chemotherapy and post-chemo/IO (E) in fast (orange) and slow (blue) progressors. (F) Phenotypic composition of CD8 T cell clonotypes that are not shared across time points. Each sample was randomly downsampled to 100 cells (if the sample had that many CD8 T cells) before visualizing overall phenotypic compositions. (G)-(J) Phenotypic distribution of newly appeared clonotypes post-chemotherapy (G), (H) and post-immunotherapy (I), (J) in slow (H), (J) and fast (G), (I) progressors. (K)-(L) Proportion of clones that are new at post-chemotherapy (K) and post-chemo/IO (L) time points in fast (orange) and slow (blue) progressors.

**Figure S3.**
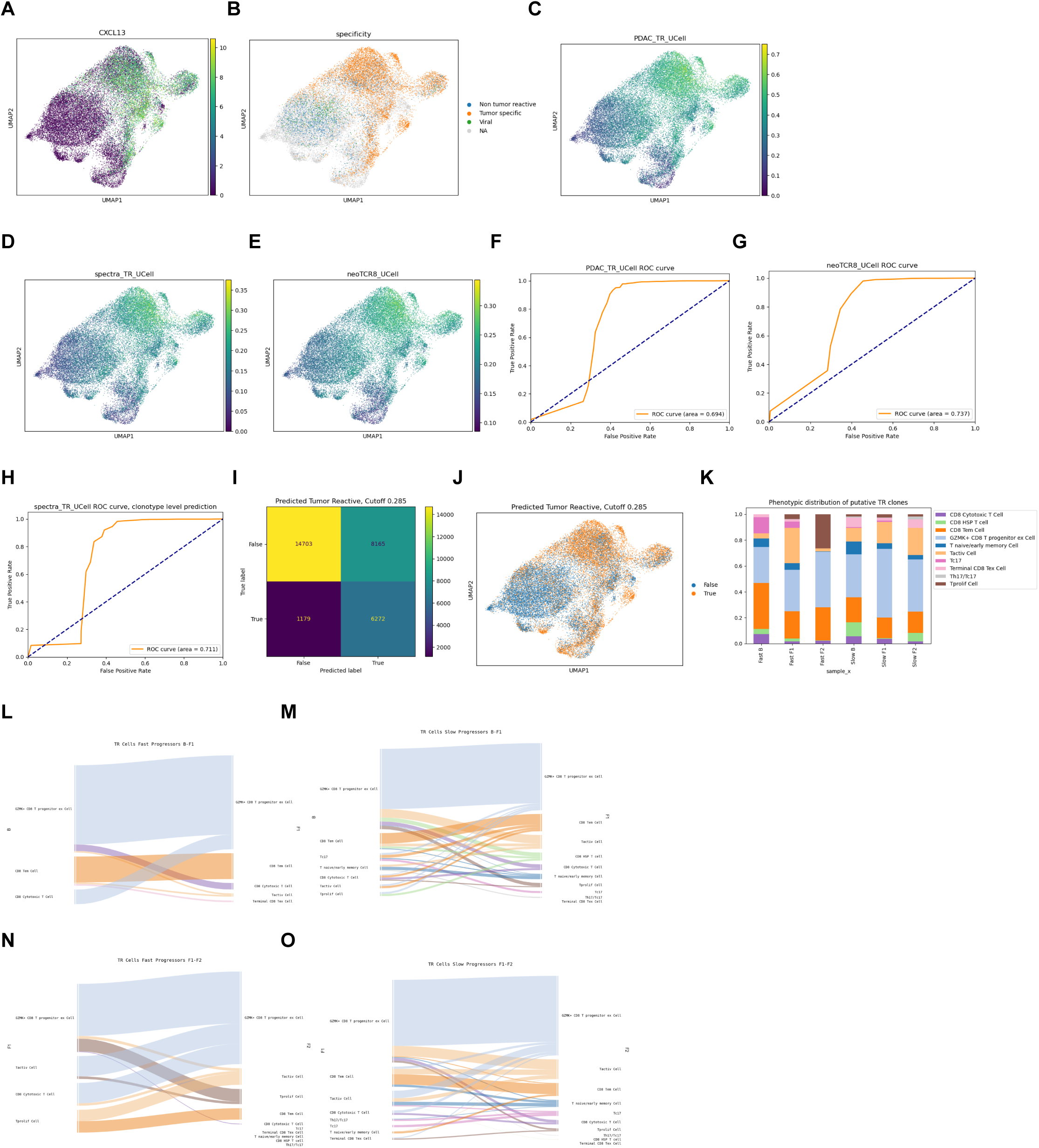
(A) UMAP representation of CXCL13 expression in T cells with paired TCR data from^32^. (B) UMAP representation of ground-truth clonal assignments of T cells with paired TCR data from^32^. (C)-(E) UMAP representation of UCell scoring on T cells with TCR data from ^32^ using genes scores from neoTCR8^10^ (C), Spectra^12^ (D), and a PDAC-specific tumor reactive signature^34^ (E). (F)-(H) AUC-ROC curves for prediction of tumor reactive clones from^32^ using UCell scoring from neoTCR8^10^ (F), Spectra^12^ (G), and a PDAC-specific tumor reactive signature^34^ (H). (I) Confusion matrix showing results of prediction of tumor reactive clones using UCell scoring with Spectra^12^ signature, with a threshold of 0.285. (J) UMAP representation of predicted tumor reactive and non-tumor reactive cells from^32^ using UCell scoring with Spectra^12^ signature, with a threshold of 0.285. (K) Phenotypic distribution of putative tumor reactive clones, averaged over patients, stratified by treatment time point and progression status. (L)-(O) Alluvial plots showing phenotypic composition of individual putative tumor reactive clonotypes shared between baseline and post-chemotherapy (L), (M) and post-chemotherapy and post-chemo/IO (N), (O) in fast (L), (N) and slow (M), (O) progressors, shown by predominant phenotype at the later (right) time point in each plot, and full proportional composition shown on earlier (left) time point in each plot. For clonotypes for which more than one phenotype was most frequent, for the purposes of visualization a predominant phenotype for that clonotype between the most frequent ones was chosen at random.

**Figure S4.**
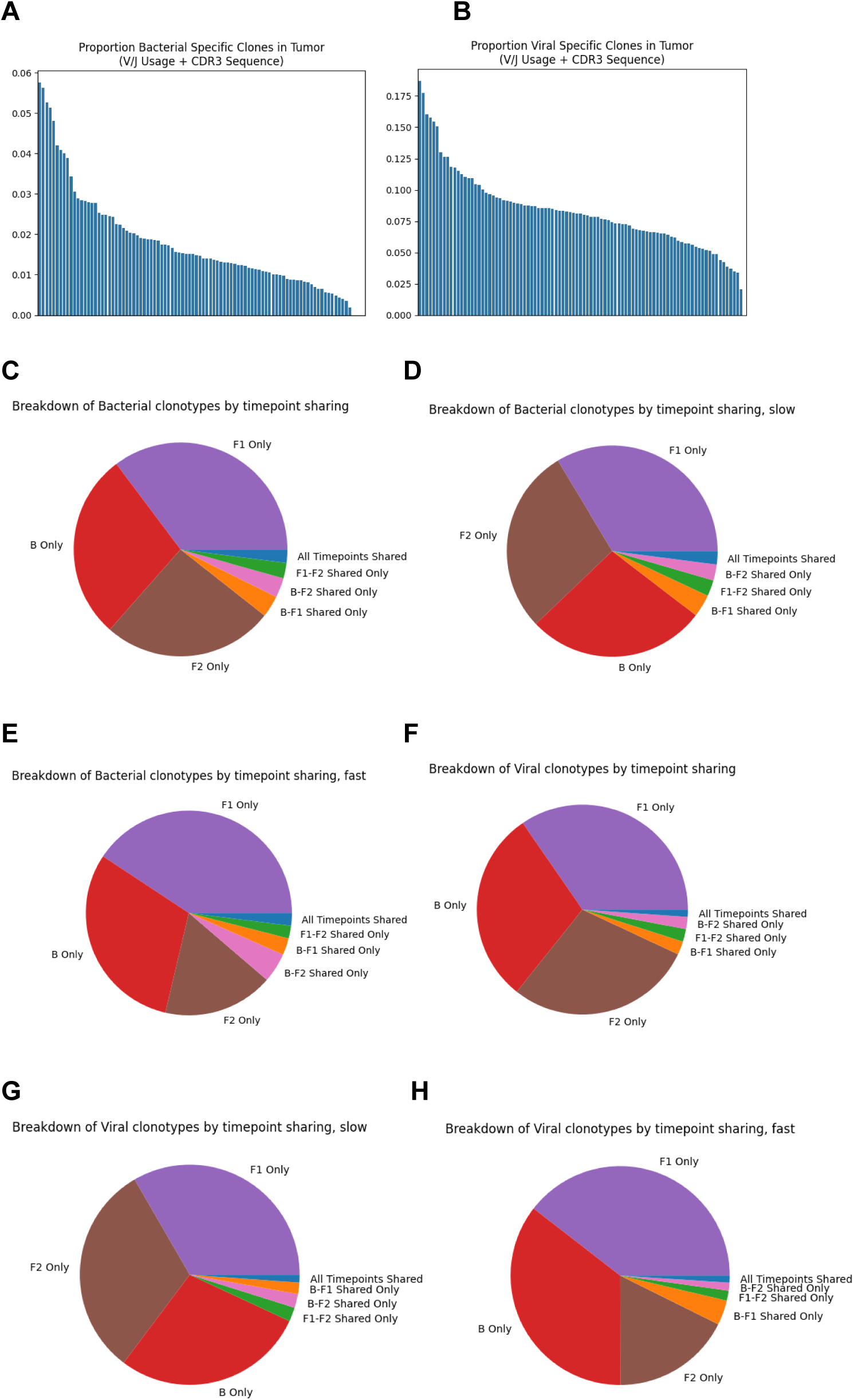
(A)-(B) Proportion of clonotypes across samples that are bacterial (A) and viral (B) reactive, according to CDR3 sequences available on public databases. (C)-(H), Pie charts showing proportion of bacterial (C) and viral (F) reactive based on the time point(s) each clonotype is present at, and split into fast (E), (H) and slow (D), (G) progressors.

**Figure S5.**
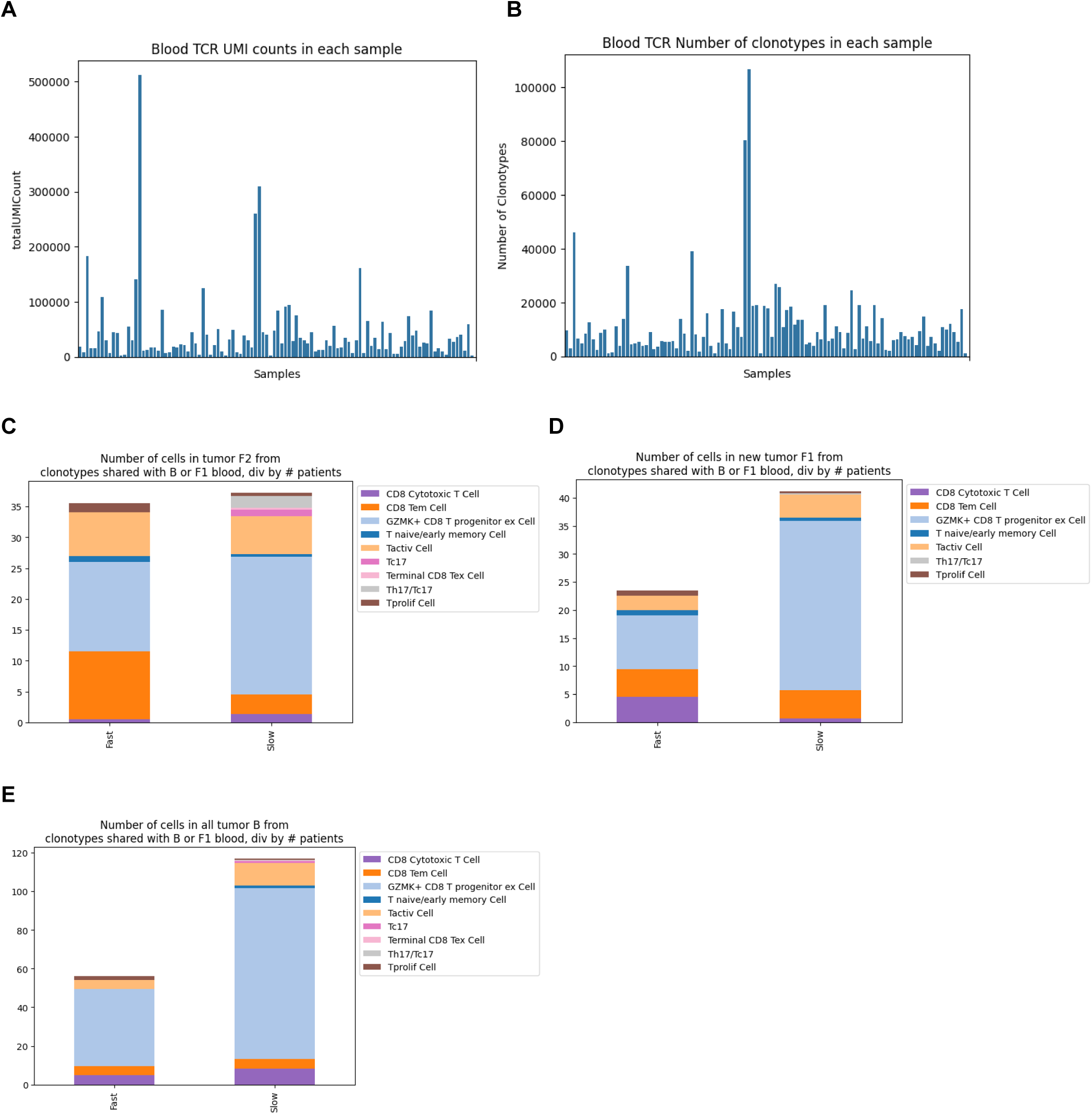
(A), (B) Number of clonotypes (A) and UMI counts (B) in each blood bulk TCR sample. (C)-(E) Number of cells divided by number of patients with shared pre-treatment and post-chemotherapy blood clonotypes that were new in the tumor post-chemo/IO (C), new in the tumor post-chemotherapy (D) and present in the tumor pre-treatment (E).

**Figure S6.**
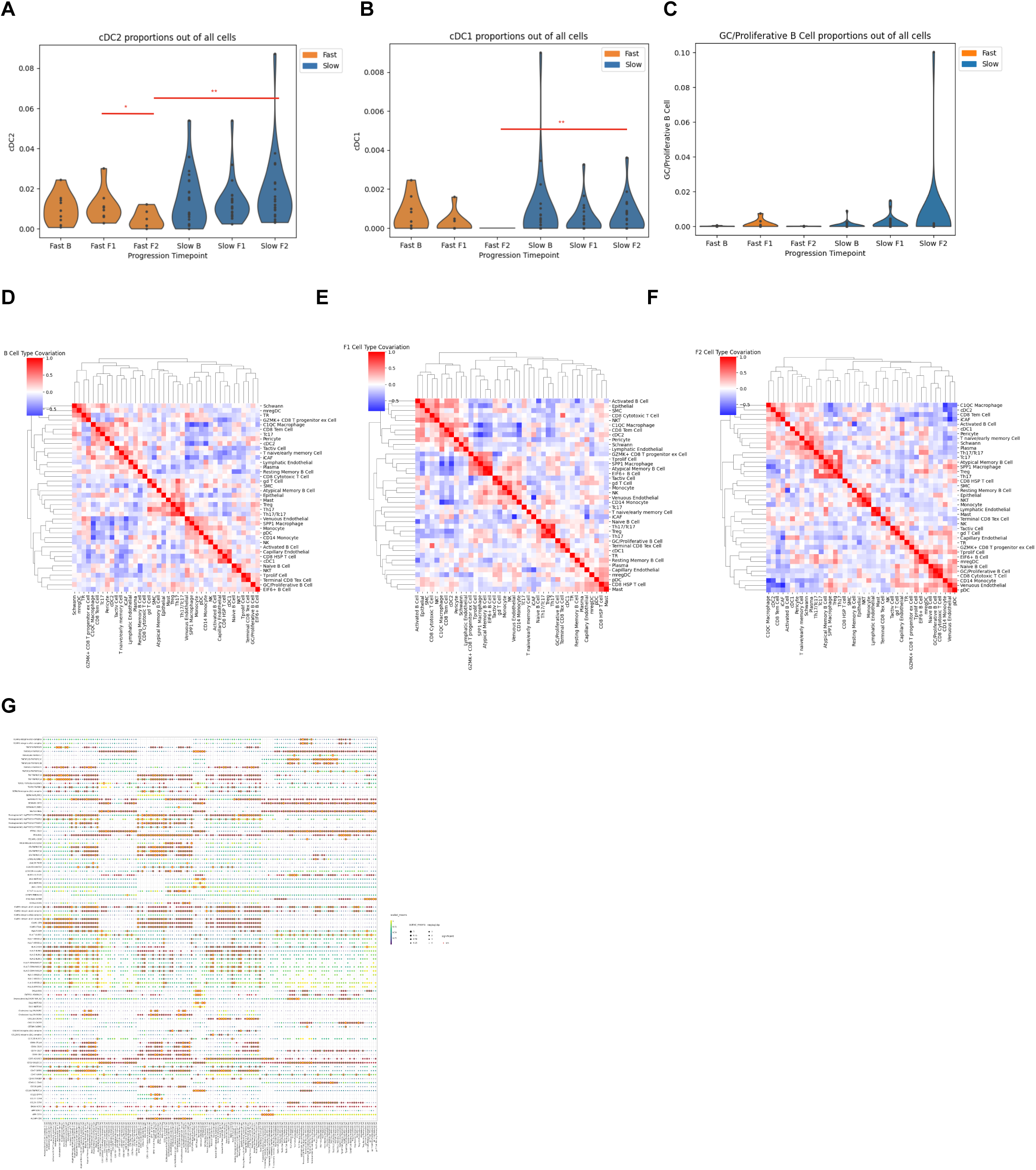
(A)-(C) Proportions of cDC1 (A), cDC2 (B) and GC/Proliferative B cells (C) in each patient out of all cells in each sample across sampling time points. Wilcoxon rank sum test, * : p < 0.1, ** : p < 0.05, *** : p < 0.01. (D)-(F) Clustered heatmap showing correlations between cell proportions in the tumor microenvironment in pre-treatment (D), post-chemotherapy (E) or post-chemo/IO tumor (F). (G) CellPhoneDB output dotplot showing significant interactions between B cells and T cells. Color indicates CellPhoneDB interaction score, size indicates the -log10(p-value) and if the interaction for a given cell type pair is significant, it is circled in red.

## References

1. Rha, S.Y., Oh, D.-Y., Yañez, P., Bai, Y., Ryu, M.-H., Lee, J., Rivera, F., Alves, G.V., Garrido, M., Shiu, K.-K., et al. (2023). Pembrolizumab plus chemotherapy versus placebo plus chemotherapy for HER2-negative advanced gastric cancer (KEYNOTE-859): a multicentre, randomised, double-blind, phase 3 trial. Lancet Oncol. 24, 1181–1195.

2. Janjigian, Y.Y., Shitara, K., Moehler, M., Garrido, M., Salman, P., Shen, L., Wyrwicz, L., Yamaguchi, K., Skoczylas, T., Campos Bragagnoli, A., et al. (2021). First-line nivolumab plus chemotherapy versus chemotherapy alone for advanced gastric, gastro-oesophageal junction, and oesophageal adenocarcinoma (CheckMate 649): a randomised, open-label, phase 3 trial. Lancet 398, 27–40.

3. Qiu, M.-Z., Oh, D.-Y., Kato, K., Arkenau, T., Tabernero, J., Correa, M.C., Zimina, A.V., Bai, Y., Shi, J., Lee, K.-W., et al. (2024). Tislelizumab plus chemotherapy versus placebo plus chemotherapy as first line treatment for advanced gastric or gastro-oesophageal junction adenocarcinoma: RATIONALE-305 randomised, double blind, phase 3 trial. BMJ 385, e078876.

4. Shitara, K., Janjigian, Y.Y., Ajani, J., Moehler, M., Yao, J., Wang, X., Chhibber, A., Pandya, D., Shen, L., Garrido, M., et al. (2025). Nivolumab plus chemotherapy or ipilimumab in gastroesophageal cancer: exploratory biomarker analyses of a randomized phase 3 trial. Nat. Med. 31, 1519–1530.

5. Kwon, M., An, M., Klempner, S.J., Lee, H., Kim, K.-M., Sa, J.K., Cho, H.J., Hong, J.Y., Lee, T., Min, Y.W., et al. (2021). Determinants of Response and Intrinsic Resistance to PD-1 Blockade in Microsatellite Instability-High Gastric Cancer. Cancer Discov. 11, 2168–2185.

6. Sun, K., Xu, R., Ma, F., Yang, N., Li, Y., Sun, X., Jin, P., Kang, W., Jia, L., Xiong, J., et al. (2022). scRNA-seq of gastric tumor shows complex intercellular interaction with an alternative T cell exhaustion trajectory. Nat. Commun. 13, 4943.

7. An, M., Mehta, A., Min, B.H., Heo, Y.J., Wright, S.J., Parikh, M., Bi, L., Lee, H., Kim, T.J., Lee, S.-Y., et al. (2024). Early Immune Remodeling Steers Clinical Response to First-Line Chemoimmunotherapy in Advanced Gastric Cancer. Cancer Discov., OF1–OF20.

8. Topp, B., Snyder, A., and Wolchok, J. (2023). RECISTv1.1 progression in oncology: Shades of gray. Cancer Cell. 10.1016/j.ccell.2023.04.012.

9. O’Rourke, C.J., Salati, M., Rae, C., Carpino, G., Leslie, H., Pea, A., Prete, M.G., Bonetti, L.R., Amato, F., Montal, R., et al. (2023). Molecular portraits of patients with intrahepatic cholangiocarcinoma who diverge as rapid progressors or long survivors on chemotherapy. Gut. 10.1136/gutjnl-2023-330748.

10. Lowery, F.J., Krishna, S., Yossef, R., Parikh, N.B., Chatani, P.D., Zacharakis, N., Parkhurst, M.R., Levin, N., Sindiri, S., Sachs, A., et al. (2022). Molecular signatures of antitumor neoantigen-reactive T cells from metastatic human cancers. Science 375, 877–884.

11. Sade-Feldman, M., Yizhak, K., Bjorgaard, S.L., Ray, J.P., de Boer, C.G., Jenkins, R.W., Lieb, D.J., Chen, J.H., Frederick, D.T., Barzily-Rokni, M., et al. (2018). Defining T cell states associated with response to checkpoint immunotherapy in melanoma. Cell 175, 998–1013.e20.

12. Kunes, R.Z., Walle, T., Land, M., Nawy, T., and Pe’er, D. (2024). Supervised discovery of interpretable gene programs from single-cell data. Nat. Biotechnol. 42, 1084–1095.

13. Fitzsimons, E., Qian, D., Enica, A., Thakkar, K., Augustine, M., Gamble, S., Reading, J.L., and Litchfield, K. (2024). A pan-cancer single-cell RNA-seq atlas of intratumoral B cells. Cancer Cell 42, 1784–1797.e4.

14. Efremova, M., Vento-Tormo, M., Teichmann, S.A., and Vento-Tormo, R. (2020). CellPhoneDB: inferring cell-cell communication from combined expression of multi-subunit ligand-receptor complexes. Nat. Protoc. 15, 1484–1506.

15. Aliazis, K., Christofides, A., Shah, R., Yeo, Y.Y., Jiang, S., Charest, A., and Boussiotis, V.A. (2025). The tumor microenvironment’s role in the response to immune checkpoint blockade. Nat. Cancer 6, 924–937.

16. Im, S.J., Hashimoto, M., Gerner, M.Y., Lee, J., Kissick, H.T., Burger, M.C., Shan, Q., Hale, J.S., Lee, J., Nasti, T.H., et al. (2016). Defining CD8+ T cells that provide the proliferative burst after PD-1 therapy. Nature 537, 417–421.

17. Siddiqui, I., Schaeuble, K., Chennupati, V., Fuertes Marraco, S.A., Calderon-Copete, S., Pais Ferreira, D., Carmona, S.J., Scarpellino, L., Gfeller, D., Pradervand, S., et al. (2019). Intratumoral Tcf1+PD-1+CD8+ T cells with stem-like properties promote tumor control in response to vaccination and checkpoint blockade immunotherapy. Immunity 50, 195–211.e10.

18. Yost, K.E., Satpathy, A.T., Wells, D.K., Qi, Y., Wang, C., Kageyama, R., McNamara, K.L., Granja, J.M., Sarin, K.Y., Brown, R.A., et al. (2019). Clonal replacement of tumor-specific T cells following PD-1 blockade. Nat. Med. 25, 1251–1259.

19. Wu, T.D., Madireddi, S., de Almeida, P.E., Banchereau, R., Chen, Y.-J.J., Chitre, A.S., Chiang, E.Y., Iftikhar, H., O’Gorman, W.E., Au-Yeung, A., et al. (2020). Peripheral T cell expansion predicts tumour infiltration and clinical response. Nature 579, 274–278.

20. Helmink, B.A., Reddy, S.M., Gao, J., Zhang, S., Basar, R., Thakur, R., Yizhak, K., Sade-Feldman, M., Blando, J., Han, G., et al. (2020). B cells and tertiary lymphoid structures promote immunotherapy response. Nature 577, 549–555.

21. Cabrita, R., Lauss, M., Sanna, A., Donia, M., Skaarup Larsen, M., Mitra, S., Johansson, I., Phung, B., Harbst, K., Vallon-Christersson, J., et al. (2020). Tertiary lymphoid structures improve immunotherapy and survival in melanoma. Nature 577, 561–565.

22. Laumont, C.M., Banville, A.C., Gilardi, M., Hollern, D.P., and Nelson, B.H. (2022). Tumour-infiltrating B cells: immunological mechanisms, clinical impact and therapeutic opportunities. Nat. Rev. Cancer 22, 414–430.

23. Cui, C., Wang, J., Fagerberg, E., Chen, P.-M., Connolly, K.A., Damo, M., Cheung, J.F., Mao, T., Askari, A.S., Chen, S., et al. (2021). Neoantigen-driven B cell and CD4 T follicular helper cell collaboration promotes anti-tumor CD8 T cell responses. Cell 184, 6101–6118.e13.

24. Lowery, F.J., Goff, S.L., Gasmi, B., Parkhurst, M.R., Ratnam, N.M., Halas, H.K., Shelton, T.E., Langhan, M.M., Bhasin, A., Dinerman, A.J., et al. (2025). Neoantigen-specific tumor-infiltrating lymphocytes in gastrointestinal cancers: a phase 2 trial. Nat. Med. 31, 1994–2003.

25. Alban, T.J., Riaz, N., Parthasarathy, P., Makarov, V., Kendall, S., Yoo, S.-K., Shah, R., Weinhold, N., Srivastava, R., Ma, X., et al. (2024). Neoantigen immunogenicity landscapes and evolution of tumor ecosystems during immunotherapy with nivolumab. Nat. Med. 30, 3209–3222.

26. Wells, D.K., van Buuren, M.M., Dang, K.K., Hubbard-Lucey, V.M., Sheehan, K.C.F., Campbell, K.M., Lamb, A., Ward, J.P., Sidney, J., Blazquez, A.B., et al. (2020). Key parameters of tumor Epitope immunogenicity revealed through a consortium approach improve neoantigen prediction. Cell 183, 818–834.e13.

27. Gayoso, A., and Shor, J. (2025). JonathanShor/DoubletDetection: doubletdetection v4.3.0.post1. Zenodo.

28. Finak, G., McDavid, A., Yajima, M., Deng, J., Gersuk, V., Shalek, A.K., Slichter, C.K., Miller, H.W., McElrath, M.J., Prlic, M., et al. (2015). MAST: a flexible statistical framework for assessing transcriptional changes and characterizing heterogeneity in single-cell RNA sequencing data. Genome Biol. 16, 278.

29. Li, S., Sun, J., Allesøe, R., Datta, K., Bao, Y., Oliveira, G., Forman, J., Jin, R., Olsen, L.R., Keskin, D.B., et al. (2019). RNase H-dependent PCR-enabled T-cell receptor sequencing for highly specific and efficient targeted sequencing of T-cell receptor mRNA for single-cell and repertoire analysis. Nat. Protoc. 14, 2571–2594.

30. Chen, S.-Y., Yue, T., Lei, Q., and Guo, A.-Y. (2021). TCRdb: a comprehensive database for T-cell receptor sequences with powerful search function. Nucleic Acids Res. 49, D468–D474.

31. Bassez, A., Vos, H., Van Dyck, L., Floris, G., Arijs, I., Desmedt, C., Boeckx, B., Vanden Bempt, M., Nevelsteen, I., Lambein, K., et al. (2021). A single-cell map of intratumoral changes during anti-PD1 treatment of patients with breast cancer. Nat. Med. 27, 820–832.

32. Oliveira, G., Stromhaug, K., Klaeger, S., Kula, T., Frederick, D.T., Le, P.M., Forman, J., Huang, T., Li, S., Zhang, W., et al. (2021). Phenotype, specificity and avidity of antitumour CD8+ T cells in melanoma. Nature 596, 119–125.

33. Andreatta, M., and Carmona, S.J. (2021). UCell: Robust and scalable single-cell gene signature scoring. Comput. Struct. Biotechnol. J. 19, 3796–3798.

34. Meng, Z., Rodriguez Ehrenfried, A., Tan, C.L., Steffens, L.K., Kehm, H., Zens, S., Lauenstein, C., Paul, A., Schwab, M., Förster, J.D., et al. (2023). Transcriptome-based identification of tumor-reactive and bystander CD8 T cell receptor clonotypes in human pancreatic cancer. Sci Transl Med 15, eadh9562.

